# A Framework for Inclusive and Accessible Clinical Research in Rare Diseases

**DOI:** 10.1101/2025.05.07.25326565

**Authors:** Lavanyaa Manjunatha, MS Saundarya, Nisha Venugopal, Jenifer Ngo Waldrop, Linda Goler Blount, Reena V Kartha, Harsha K Rajasimha

## Abstract

**Background:** Equitable representation of all populations is crucial for generalizing rare disease (RD) clinical research outcomes, especially given the low prevalence and geographically sparse distribution of patients with RDs. In our companion manuscript (*Current State and Demographic Trends of Medically Underserved Populations in Rare Disease Research*, Manjunatha *et al.*) we quantified reporting of demographics, socioeconomic factors (SF), and participation trends of medically underserved populations (MUPs) in RD research and clinical trials.

**Methods:** The study builds on the findings of Manjunatha *et al*., where we analyzed the reporting of demographics and SF in RD clinical research, here we perform a representation and policy gap analysis of this extracted data. The representation analysis evaluated 13 variables, including age, sex or gender, race, ethnicity, and SF, using four key analyses: reporting statistics, representation, participant distribution, and benchmarking against the US census data. The qualitative policy analysis included existing national and international policies and guidelines.

**Results:** Only age, sex or gender, race, and ethnicity had sufficient data for the representation analysis. While diversity was moderate for these variables, equity, inclusion, and accessibility were low, particularly for racial and ethnic minorities, nonbinary genders, and older adults. Data were insufficient for MUPs such as lesbian, gay, bisexual, transgender, queer or questioning individuals, rural residents, veterans, military spouses, people affected by poverty, and religious minorities. Based on the representation analysis and building upon existing foundational policies and guidelines, we propose three recommendations and a six-pillar framework to mandate and standardize data reporting practices and improve the representation of MUPs in RD clinical research with broader relevance to all clinical research in general. The six pillars are patient advocacy, policy legislation, governmental oversight, standardized data collection and reporting, technological enablement, and global epidemiological research.

**Conclusions:** Addressing the historical underrepresentation of MUPs requires upgrading the foundation of clinical research instead of a piecemeal, siloed approach. This study underscores the systemic gaps in the representation of MUPs in RD research and proposes a six-pillar actionable framework to address these disparities. The systematic implementation of these six pillars can enhance the integrity and outcomes of future RD clinical research.

## Introduction

Equitable representation of all populations, including medically underserved populations (MUPs) in clinical research is crucial to ensure the generalizability of the evidence, maintain public trust in the healthcare industry, and unlock medical innovation and the full potential of precision medicine to improve health outcomes for all (1). Diversity, equity, inclusion, and accessibility (DEIA) is critical in rare diseases (RDs), given their low prevalence and sparse geographical distribution of affected individuals. Except for a few studies, little is known about the comprehensive MUPs in RD clinical trials (2–5).

Our companion manuscript (6) addressed this issue by analyzing the reporting of and participant distribution in 13 demographic and socioeconomic factors (SFs) in RD clinical research in the United States (US) over 40 years. Manjunatha *et al.* assessed MUPs, including children, older adults, women, lesbian, gay, bisexual, transgender, queer or questioning (LGBTQ+) individuals, historically underserved racial and ethnic groups, multiracial groups, rural residents, veterans, religious minorities, immigrants, people living with disabilities, and those disadvantaged by poverty. The key findings were the underreporting of race, ethnicity, and SF; the lack of standardized reporting in publications; and the underrepresentation of MUPs. Historically underserved racial and ethnic groups were underrepresented in RD trials compared with the US census. There was little or no data on LGBTQ+ individuals, members of religious groups, rural residents, veterans, people living with disabilities, and those disadvantaged by poverty.

Recent recommendations or frameworks to improve clinical trial diversity include the American Society of Hematology (ASH) DEI toolkit, Diversity Convergence Project (DCP), the Multi-Regional Clinical Trials (MRCT) Center’s Achieving Diversity, Inclusion, and Equity in Clinical Research guidance and toolkit, and recommendations from the National Academy of Science, Engineering, and Medicine (NASEM) (7–10). RD patient advocacy groups and researchers have underscored the need to enhance DEIA to achieve health equity (11,12). The proposed solutions range from innovative trial designs, increasing patient participation in guiding trial design, including logistical considerations, standardized reporting of population data in electronic health records (EHRs), enhancing cultural competency, engaging with diverse communities, and routine monitoring and evaluation of DEIA initiatives (11–13). These frameworks and solutions provide recommendations in a silo without duly considering how they may overlap with existing foundational national and international policies and guidelines for clinical research, particularly for RDs.

Our study aimed to develop an actionable framework to improve the representation of MUPs in RD research. Specifically, we 1) evaluated the gaps in MUP representation and 2) analyzed relevant national and international policies or legislative documents governing clinical research. Accordingly, we developed three recommendations and a six-pillar framework.

## Methods

### Representation analysis

We performed a representation analysis of the results reported in our companion manuscript (6). Data for 13 demographics and SF variables and their subcategories were extracted and evaluated based on 1) reporting statistics, 2) representation, 3) participant distribution statistics, and 4) comparing participant distribution with US census data as a benchmark (Additional File 1: Table S1). See Additional File 2: Supplementary methods for details. Diversity was evaluated based on the reporting statistics and representation. Equity and inclusion were evaluated based on distribution statistics and comparisons with the US census as an external benchmark. Accessibility was inferred from assessments of diversity, equity, and inclusion, as reporting and distribution statistics cannot serve as direct markers for how trials were made accessible.

### Policy analysis

The Food and Drug Administration (FDA), National Institutes of Health (NIH), ClinicalTrials.gov, Congress.gov, and the US Department of Health and Human Services websites were searched for policies, guidance documents, laws, bills, and guidelines related to clinical research published up to 2024. Google searches were performed using specific keywords (Additional File 2: Supplementary methods). The documents (hereafter referred to as policy) were evaluated to identify if they reported 1) measures of diversity, 2) provisions for increasing diversity, and 3) recommendations or frameworks for improving inclusion and accessibility in clinical trials.

## Results

### Representation and policy gaps analysis

The representation analysis (Table 1) revealed several gaps in RD clinical research, which we illustrate using age as an example. The age variable was scored “high” along the diversity axis as it was reported in >75% of publications and clinical trials and had data for subcategories (child; adult; older adult; child and adult; child, adult, and older adult; adult and older adult). However, diversity for age subcategories “child” and “older adults” was scored “low” because of the lower reporting levels. Children and older adults were scored “low” for inclusion based on the low participant proportions in trials. Similarly, equity was scored “low” based on participant distribution alone rather than a direct comparison with the US census, as three of the six age subcategories included mixed age groups, making it difficult to make meaningful comparisons. Accessibility was also inferred to be “low” for older adults and children based on diversity, equity, and inclusion assessments. The overall assessments of the 13 variables are detailed in Additional File 1: Table 1.

**Table 1:**
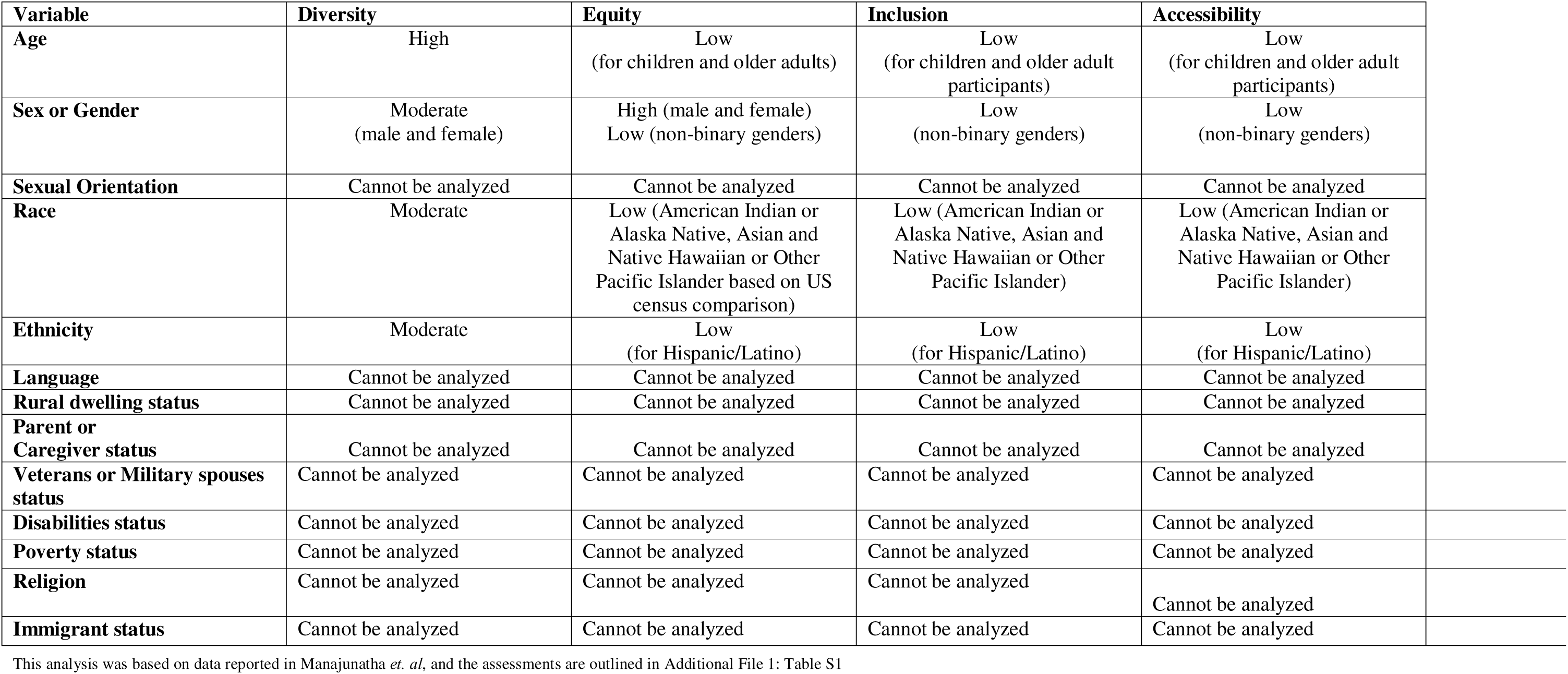
Evidence-based categorical assessment of Diversity, Equity, Inclusion, and Accessibility in US-based RD clinical research.

Of the different variables, only age, sex or gender, race, and ethnicity could be evaluated for DEIA due to reporting levels and the availability of granular data (Table 1, eTable1). Diversity was assessed as “high” for age and moderate for sex or gender, race, and ethnicity based on the reporting statistics and categorical diversity or representation. However, equity, inclusion, and accessibility were assessed as “low” for these characteristics. For male, female, and White participants, equity, inclusion, and accessibility were assessed to be “high.” In contrast, they were “low” for older adults, nonbinary gender, American Indian or Alaska Native, Asian, Black or African American, and Native Hawaiian or Other Pacific Islander participants. Representation analysis was possible for only 4 of the 13 variables because of the significant lack of information reporting on several variables, including SF. Table 2 provides an overview of the national and international policies, laws, and guidelines included in the policy analysis and their current scope. Age, sex, race, and ethnicity were the most common measures of diversity mentioned in most of these documents, which aligned with our analysis (Table 1). Only the 21^st^ Century Cures Act focuses on improving the inclusion of sexual minorities. The Food and Drug Omnibus Reform Act of 2022 (FDORA) and the resulting FDA draft guidance (2024) acknowledged SF as variables affecting trial diversity (14–16). Table 2 also presents the proposed potential policy enhancements to improve the scope of the existing provisions to improve MUP participation.

**Table 2:** Foundational and recent policies^a^ governing clinical research^b^.

| Policy | Current DEIA focus or scope of the policy | Suggested enhancements for incorporating DEIA principles |
| --- | --- | --- |
| The Belmont Report (44) | The Belmont Report is a statement of ethical principles and guidelines for the protection of human subjects of research. It covers the three basic ethical principles of respect for persons, beneficence, and justice. The broad applications to clinical research are:<br>• informed consent – information, comprehension, and voluntariness<br>• risks and benefits – at the individual and societal level<br>• selection of subjects – fair selection of subjects, without exploitation | The foundational ethical principles of the Belmont Report are broadly aligned with the principles of DEIA. It can benefit from explicitly mentioning inclusion and accessibility in the principles and applications.<br>• For instance, the “respect for person” principle can be updated to include non-binary genders and other MUPs such as rural dwellers, veterans, caregivers, immigrants, religious minorities, and other historically underserved groups. |
| The Helsinki Declaration (43),<br>Original: 1964, Latest revision: 2024 | The Helsinki Declaration is a statement of ethical principles for medical research involving human participants covering a range of topics, including:<br>• confidentiality – maintaining the privacy of participants<br>• informed consent – inform about aims, methods, risks, and benefits in plain language; right to refuse or withdraw consent; plain language<br>• vulnerable population | • Privacy protection assurances take into consideration data that could be used to stigmatize or discriminate against, such as SOGI data<br>• Consent forms and other materials should be made available in all relevant languages based on the participant recruitment<br>• Consent forms and other materials should be accessible to people living with disability and older adults<br>• Clearly specify who vulnerable populations are |
| The Orphan Drug Act (21) 1983 | The law encourages pharmaceutical companies to develop therapies for rare disease by providing:<br>• amendments to Federal Food, Drug, and Cosmetic Act<br>• recommendations for investigations of drugs for rare diseases or conditions<br>• designation of drugs for rare diseases or conditions<br>• Protection for unpatented drugs for rare diseases or conditions<br>• open protocols for investigations of drugs for rare diseases or conditions<br>• grants and contracts for development of drugs for rare diseases and conditions<br>• orphan products board<br>• annual report | Revise the law to include the following sections:<br>• standardized data collection practices for demographics, including SOGI, SF, and special status |
| The International Council for Harmonisation (ICH) Harmonised Guideline Good Clinical Practice (GCP) E6 (R3) (45), Original: 1996, revised draft: 2023 | ICH GCP guidelines aim to provide a unified standard to facilitate the mutual acceptance of clinical trial data for ICH member countries and regions by applicable regulatory authorities.<br>The guidelines cover the following three broad areas that can influence diversity in clinical trials:<br>• informed consent: clear and concise language, well-informed, voluntary participation<br>• sponsor: trial design, quality, and oversight, data records<br>• data governance: safeguarding data integrity, confidentiality, and security | • Consent forms and other materials should be made available in all relevant languages based on the participant recruitment<br>• Consent forms and other materials should be accessible to people living with disability and older adults<br>• Require workforce training and cultural competency for investigators, sponsors, and researchers to work with a diverse participant group<br>• Require standardized collecting of demographics, including SOGI, SF, and special status data<br>• Require sponsors to have enrolment plans to recruit diverse populations |
| Common Rule (46),<br>(45 CFR 46), Original: 1991,<br>Revised: 2018 | The 45 CFR 46 is a blanket federal policy for the protection of human subjects and provides guidance on:<br>• equitable selection of research subjects<br>• protection of rights and welfare of vulnerable research subjects, including children, prisoners, individuals with impaired decision-making capacity, or economic or educationally disadvantaged persons<br>• informed consent – information should be given in understandable language | The 45 CFR 46 can be enhanced by:<br>• expanding to include diverse and inclusive subpopulation recruitment in guidance for research subjects<br>• clarifying that informed consent should be made available in different languages based on the participant’s spoken language and should also consider accessibility requirements |
| NIH Revitalization Act of 1993, (47), 1993 | The legislation emphasized the inclusion of women and underrepresented racial and ethnic minorities clinical research by:<br>• mandating their inclusion in all NIH-funded clinical research unless scientifically or ethically inappropriate<br>• requiring subgroup analysis to identify any treatment differences in these groups<br>• improving the representation of women physicians and scientists conducting research in national institutes<br>• creation of a national data system and clearinghouse on research on women’s health to collect and disseminate data on women’s health including age, sex, race, ethnicity and location of trial sites<br>• biennial reporting by the NIH Advisory Committee | The act can be enhanced by expanding the reporting to include:<br>☐ SOGI, SF, and special status |
| Health Insurance Portability and Accountability Act of 1996 (48) (HIPAA), 1996<br>HIPAA Privacy Rule<br>HIPAA Security Rule | The HIPAA Act establishes federal standards protecting sensitive health information from disclosure without a patient's consent and has a strong bearing on informed consent in clinical trials. | Although HIPAA does not directly address clinical trials, as HIPAA is followed when obtaining informed consent, following these principles can enhance DEIA:<br>• accessible consent forms that consider the needs of diverse populations, such as people from varied linguistic backgrounds, children, and those with disabilities<br>• privacy protection assurances take into consideration data that could be used to stigmatize or discriminate against such as SOGI data |
| FDA Amendments Act of 2007 (49) (FDAAA 801 and the Final Rule 2017) | The FDAAA 801 and Final rule aims to improve clinical trial transparency by requiring demographics, baseline data, outcomes, and adverse events data to be reported on ClinicalTrials.gov. | ● Expand the demographics reporting requirements to include SOGI<br>● Expand the data reporting to include SF and special status |
| Food and Drug Administration Safety and Innovation Act (23) (FDASIA), 2012 | The FDASIA Act has provisions for ensuring the inclusion of demographic data (age, sex, race, and ethnicity) in clinical trials and data analysis in applications for drugs, biologics, and devices. | The safety and efficacy by subpopulation analysis can be expanded to include SOGI and SF in applications for drugs, biologics, and devices to help understand how these measures of diversity affect health outcomes. |
| 21st Century Cures Act (14) 2016 | The 21<sup>st</sup> Century Cures Act aims to accelerate medical product development and bring innovations to patients. Several sections, including 2038, 2039, 404M, 404N, and 498, are relevant to diversity and inclusion in clinical trials, specifically for:<br>● women, members of minority groups, relevant age categories, sexual and gender minority populations<br>law focuses on:<br>● diversity of populations studied for clinical research with respect to biological, social, and other determinants of health contributing to disparities<br>● circumstances for reporting and analyzing data, including disaggregation by disease condition and demographics<br>● data and information sharing in accordance with privacy laws and regulations<br>● promoting workforce diversity<br>Real-world evidence – data collection standards and methodologies | The 21st Century Cures Act broadly covers all MUPs; however, the diversity scope can be enhanced by:<br>● explicitly including MUPs such as caregivers, veterans, immigrants, and religious minorities.<br>● mandating for standardized data collection and reporting practices<br>● including additional privacy assurances for sensitive data that can be used to stigmatize patients, such as SOGI |
| NIH Inclusion Across Lifespan Policy (50), 2017 | The policy focuses on including individuals of all ages in all NIH-supported research.<br>Applications and proposals are required to:<br>• describe plans for individuals across the lifespan<br>• provide the rationale for selecting participants of a specific age range<br>• submit a progress report with deidentified participant data for age, sex/gender, race, and ethnicity to NIH | The Inclusion Across Lifespan Policy can be enhanced to expand the participant data reporting to include SOGI, SF, and special status |
| FDA Reauthorization Act of 2017(51) (FDARA), 2017 | The FDARA covers patient access to investigational drugs by including provisions for modifying inclusion and exclusion criteria so that diverse populations can access clinical trials:<br>• infants and children<br>• pregnant and lactating women<br>• seniors<br>• individuals with advanced disease<br>• and individuals with co-morbid conditions<br>The law also mentions regulatory, geographical, and socioeconomic barriers to participation in clinical trials | The FDARA Act can be enhanced by including provisions for modifying inclusion and exclusion criteria such as individuals from diverse linguistic backgrounds and limited English proficiency can access clinical trials |
| Enhancing the Diversity of Clinical Trial Populations — Eligibility Criteria, Enrollment Practices, and Trial Designs (52) (Final Guidance), 2020 | This guidance fulfils the mandate under the section 610(a)(3) of FDARA.<br>The guidance emphasizes diversity in terms of:<br>• demographic characteristics: age, sex, race, ethnicity, and residence location<br>• non-demographic factors: organ dysfunction, comorbid conditions, disabilities, and rare disease populations | The guidance can be enhanced by including the following as measures of diversity:<br>☐ SOGI<br>☐ SF<br>☐ special status |
| Food and Drug Omnibus Reform Act of 2022 (16) (FDORA), 2022 | The FDORA has provisions to improve clinical trial diversity through:<br>• diversity action plans (DAP) for clinical studies<br>• FDA evaluation and mandate for post-market surveillance data when demographics subgroup data are insufficient<br>• public workshops to get feedback from stakeholders for increasing the enrolment of historically underrepresented populations in trials<br>• mandated annual summary reports of progress in clinical trial diversity<br>DAPs should cover enrolment:<br>• goals disaggregated by age group, sex, race, geographic location, socioeconomic status, and ethnicity.<br>• barriers such as population size, geographic location, and SF<br>The statute also covers guidance on decentralized trials using RWE, innovative trial designs, and data collection methodologies. | The FDORA Act covers diversity almost comprehensively; however, there is scope for improvement by:<br>• including SOGI data<br>• clarifying what SF data to collect and report<br>• mandating that demographics, SF, and special status data be collected and reported in a standardized manner |
| The DEPICT Act (22), 2022 | This legislation primarily emphasizes age, sex, race, and ethnicity through:<br>• annual report on progress to FDA<br>• community engagement and outreach to improve underrepresented minorities<br>• grants to increase the capacity of community health centers to participate in clinical trials<br>It also acknowledges the potential of real-world evidence (RWE) to complement traditional trial data and encourages its incorporation into trial frameworks. | Expand diversity to include:<br>• SOGI<br>• SF<br>• special status |
| The NIH Clinical Trial Diversity Act of 2023 (53) | The Bill sets out requirements to increase the diversity of clinical trial participants and requires other activities to foster participation in clinical trials.<br>Sponsors are required to provide:<br>● goals for recruiting and retaining trial participants by age, sex, race, and ethnicity, either by disease prevalence or by US population<br>● plans for achieving the recruitment and retention goals by implementing less-burdensome clinical trial follow-up requirements<br>● plans for making the trials accessible, like using fewer follow-up visits, using wearable technology, online follow-ups, and virtual telemedicine, among others<br>● annual reports of demographic data in clinical trial participants by the sponsor to NIH and subsequent publication on the NIH website<br>Additionally, researchers and other individuals involved with the trial must complete education and training about diversity in clinical trials | The Bill actively mentions diversity in the context of age, sex, race, and ethnicity and also mentions underserved populations. However, it can further be enhanced by explicitly including these axes when considering diversity in participants:<br>● SOGI<br>● SF<br>● special status |
| Postmarketing Approaches to Obtain Data on Populations Underrepresented in Clinical Trials for Drugs and Biological Products (54), 2023 | This guidance describes FDA requirements and provides recommendations for obtaining safety and effectiveness information on drugs, when appropriate, in the postmarketing setting in historically underrepresented patient populations in clinical trials. The guidance includes mechanisms to improve diversity through:<br>• Postmarketing requirements<br>• Postmarketing commitments<br>• Use of real-world data<br>The guidance recommends:<br>• submitting a diversity plan as outlined in the draft guidance *Diversity Plans to Improve Enrollment of Participants from Underrepresented Racial and Ethnic Populations in Clinical Trials* using the guidance for industry, *Enhancing the Diversity of Clinical Trial Populations Eligibility Criteria, Enrollment Practices, and Trial Designs* (November 2020) to develop recruitment strategies to enrol diverse participants | The guidance can be enhanced by expanding subpopulation analysis to also include:<br>☐ SOGI<br>☐ SF<br>☐ special status |
| Diversity Action Plans to Improve Enrollment of Participants from Underrepresented Populations in Clinical Studies (55) (Draft Guidance for Industry), 2024 | The draft guidance fulfills the requirement outlined in section 3602 (a) of FDORA.<br>According to the guidance, Diversity Action Plans (DAP) must include the sponsor's:<br>● enrollment goal disaggregated by age, sex, race, and ethnicity<br>● rationale for such goals<br>● explanation of how the sponsor intends to meet such goals.<br>The guidance also notes that sponsors should consider other factors that affect clinical outcomes, such as:<br>● socioeconomic status<br>● geographic location<br>● comorbidities<br>The guidance also acknowledges broader issues of health disparities that occur due to:<br>● geographic location<br>● gender identity<br>● sexual orientation<br>● socioeconomic status<br>● physical and mental disabilities<br>● pregnancy and lactation status<br>● co-morbidities | The draft guidance for Diversity Action Plans can be enhanced by:<br>☐ expanding the enrollment goal to include SOGI, SF, special status data<br>☐ clarifying what socioeconomic status data to collect and report<br>☐ requiring demographics, SF, special status data to be collected and reported in a standardized manner |
| Collection of Race and Ethnicity Data in Clinical Trials and Clinical Studies for FDA-Regulated Medical Products (56), 2024 | The guidance provides recommendations on:<br>● meeting the requirements of the Demographic Rule for INDs and NDAs<br>● collecting of race and ethnicity data in BLAs and medical device applications<br>● improving the completeness and quality of demographic data collection and reporting according to FDASIA Section 907 Action Plan<br>The guidance recommends collecting race and ethnicity data as per the OMB Policy Directive 15 and has provisions for using more detailed racial and ethnic categories. | The guidance can be enhanced by:<br>☐ expanding the granularity of race and ethnicity data collected as outlined in the MRCT Centre's toolkit (57)<br>☐ include guidance other demographic variables such as sex, gender, and SOGI |
| Clinical Trial Modernization Act (58), 2024 | This bill aims to improve clinical trial participation of underrepresented populations through:<br>• community education, outreach, and recruitment activities for trials<br>• payment of study participant clinical trial expenses, such as travel, transportation, and meal expenses to all participants so that participants from all relevant demographic and socioeconomic populations and geographic locations, including rural locations<br>• providing free digital health technologies to facilitate participation of underrepresented populations<br>• making trials accessible through support of clinical trial cost-sharing<br>The bill follows the underrepresented populations definition according to the National Institutes of Health for purposes of Toolkit for Patient-Focused Therapy Development. | The bill can be enhanced by expanding the underrepresented populations to include:<br>☐ sexual orientation<br>☐ SF<br>☐ special status |
| Decentralized Trials Guidance (59), 2024 | This guidance provides recommendations for sponsors, investigators, and other stakeholders in implementing decentralized elements in clinical trials such as:<br>☐ decentralized trial design and conduct<br>☐ remote control trial visits and activities<br>☐ digital health technologies<br>☐ roles and responsibilities of sponsors, and investigators<br>☐ FDA and IRB oversight<br>☐ investigational products and their packaging and shipping<br>☐ safety monitoring<br>☐ electronic systems used for conducting decentralised trials | In the roles and responsibility section for sponsors, the guidance can be enhanced by including the following as measures of diversity:<br>☐ SOGI<br>☐ SF<br>☐ special status<br>The informed consent and the IRB oversight section can be enhanced to include guidance for electronic consent forms and materials to be:<br>☐ in relevant languages for the trial participants<br>☐ made accessible for a diverse participant group |

<sup>a</sup>Includes policies, guidance documents, bills, laws, and guidelines concerning clinical research and rare diseases
<sup>b</sup>Ordered based on the chronology of the release date of the original documents.
Special status includes veteran, caregiver, disability, immigrant, and religion
Abbreviations: BLA, Biologics License Application; DEIA, diversity, equity, inclusion, and accessibility; IND, Investigation New Drug; IRB, Institutional Review Boards; MUPs, medically underserved populations; SF, socioeconomic factors; SOGI, sexual orientation and gender identity; RWE, real-world evidence,

### Recommendations and Framework for improving MUP representation in RD clinical research

Our companion manuscript (6) and analysis revealed a lack of standardized data reporting, inclusion, and accessibility of MUPs in RD research. To address these gaps, we recommend the following: 1) complete, clear, accurate, and standardized collection and reporting of demographics, including sexual orientation, gender identity (SOGI), SF, and special status, such as veteran, caregiver, disability, immigrant, and religion in publications and clinical trials; 2) remove barriers to including MUPs in clinical trials; and 3) enhance the accessibility of trials for increased MUP participation. Table 3 elaborates on these recommendations and provides a rationale. We propose an actionable six-pillar framework (Figure 1) to apply these three recommendations for enhancing DEIA in RD clinical research by drawing on our evidence-based assessment (Table 1, Additional File 1: Table S1) and analyzing gaps in existing policies, laws, and guidelines (Table 2).

**Figure 1:**
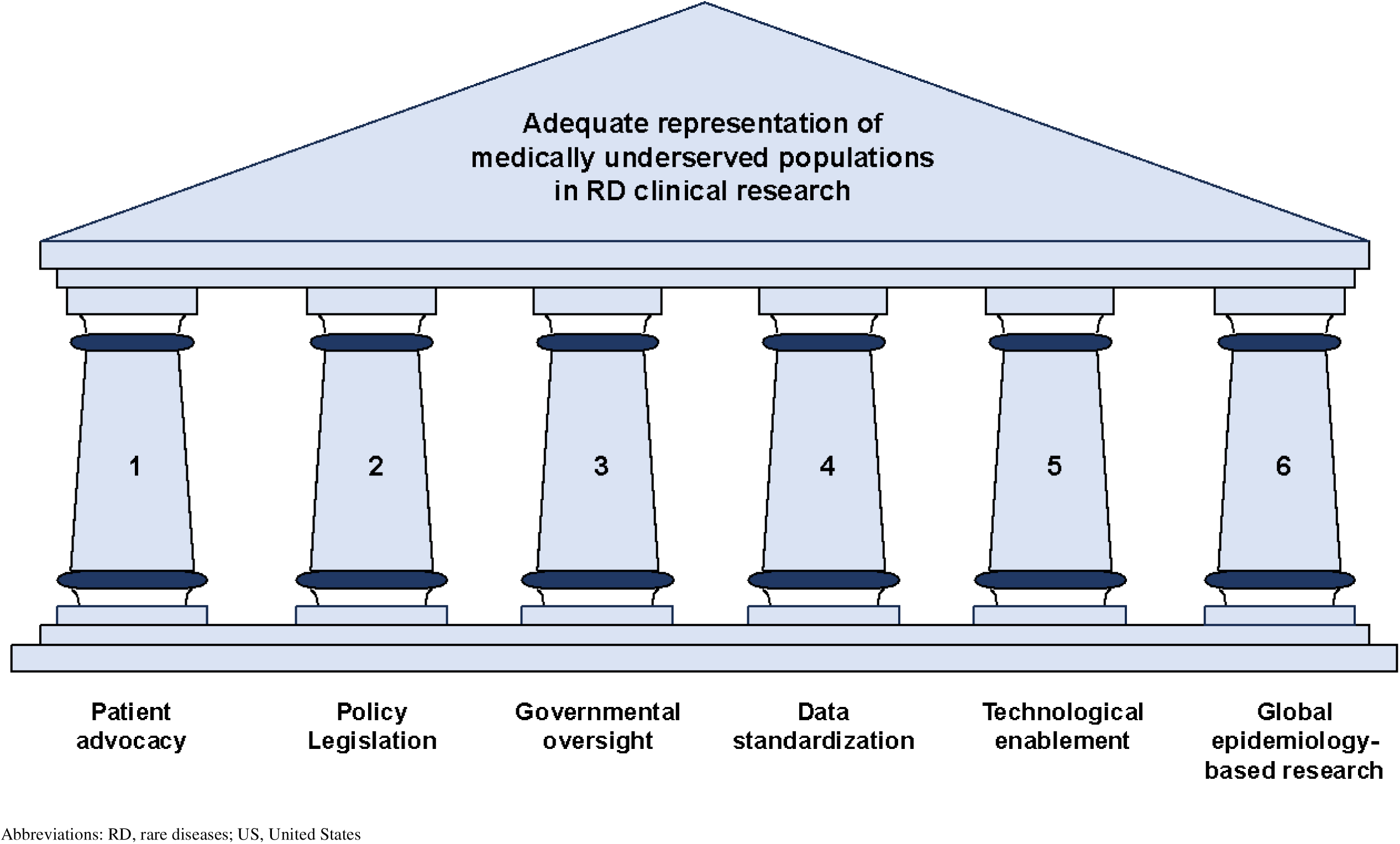
Framework for improving the representation of medically underserved populations in US-based RD clinical research.

**Table 3:** Recommendations^a^ for improving DEIA in publications and clinical trials.

| Recommendations | Rationale |
| --- | --- |
| **Complete, clear, accurate, and standardized collection and reporting of demographics, SF and special status data in publications and clinical trials**<br>1. **Sex or gender data:**<br>a. Clarify whether the reported data are for sex or gender in publications and clinical trials<br>2. **SOGI data:**<br>a. For sexual orientation: lesbian or gay, bisexual<br>b. For gender identity: male, female, trans-male, trans-female, non-binary or gender non-conforming, different identity, choose not to respond<br>3. **Race and Ethnicity**<br>a. Use NIH/OMB format for reporting race and ethnicity data in publications<br>b. Increase the collection and reporting of details of race and ethnicity subcategories in clinical trial data<br>o For example, increase the granularity of data collected for the Asian race category to include all subpopulations possible<br>c. Report both aggregated and disaggregated data for all race and ethnicity subcategories in publications and clinical trials<br>4. **Language:**<br>a. Require publications to report if language was used as an inclusion or exclusion criterion for all reports on clinical trials<br>5. **SF data:**<br>a. Make them standardized data elements in clinical trials' baseline characteristics<br>b. Collect and report language spoken, education, geographical location, income, insurance<br>c. Geographic location: expand the region of enrollment data element in clinical trials to include details about where the participants were recruited from. For example, details such as how far was the trial center from their residence or if they lived in a rural, metro, or urban area<br>6. **Special status:**<br>a. Veteran, caregiver, disability and immigration status, and religion | Our recommendations are aimed at improving transparency and accountability in clinical research, achieving equity in representation, and data harmonization for greater interoperability such that future DEIA efforts in clinical research and policy can be data and evidence-driven. |
| **Remove barriers to including MUPs in clinical trials**<br>1. Remove language-based inclusion/exclusion criteria in clinical trials<br>2. Engage with patient advocacy groups to understand the needs of MUPs such as LGBTQ+ individuals, older adults, etc<br>3. Relaxation of inclusion/exclusion criteria for older adult participants | To improve the participation of MUPs |
| **Enhance the accessibility for MUPs to participate in clinical trials** | To make RD trials more accessible for people from varied linguistic backgrounds, older adults, those affected by disability, having caregiving responsibilities, affected by poverty, and living in remote locations |
| 1. Make recruitment and patient materials available in relevant languages for the target recruitment population<br>2. Use decentralized trials, virtual trials, wearable technology |  |

<sup>a</sup>Based on the representation analysis (Table 1) of data presented in the companion manuscript (*Manjunatha et al.*).
Abbreviations: DEIA, diversity, equity, inclusion, and accessibility; FDA Food and Drug Administration; LGBTQ+, lesbian, gay, bisexual, transgender, queer or questioning, MUPs, medically underserved populations, NIH/OMB, National Institute of Health/Office of Management and Budget; RD, rare disease; SF, socioeconomic factors; SOGI, sexual orientation and gender identity

#### Pillar 1: Patient advocacy

Patient advocacy is the hallmark of progress in RD clinical research. Patient advocacy groups (PAGs) support research, create patient registries, conduct natural history studies, contribute to clinical trials, and educate healthcare providers and patients. PAGs can play a crucial role in enhancing MUP participation in RD clinical research by 1) understanding and advocating the needs of the affected populations; 2) incorporating patient feedback to remove barriers to inclusivity, such as native languages; 3) facilitating trust and engagement by providing culturally competent educational materials to stakeholders; 4) advocating for comprehensive DEIA reporting and moral policing in RD research; and 5) forming PAG coalitions to advocate for policy amendments to the bilateral and bicameral RD caucus.

#### Pillar 2: Policy legislation

Legislative advocacy is required to adopt and mandate the implementation of new guidelines for improving MUPs’ representation in RD clinical research. We recommend1) revising existing bills or laws such as the Orphan Drug Act of 1983, 21^st^ Century Cures Act, DEPICT Act of 2002, and NIH Clinical Trial Diversity Act of 2023, to include SOGI and SF data as measures of diversity; 2) adapting the social determinants of health-related International Classification of Diseases (ICD)-10 Z codes from Centres for Medicare & Medicaid Services (17) data to enhance collection and reporting SF data in clinical research; 3) mandating expedited implementation of ICD-11 (18,19) to better track RDs; 4) modernizing newborn screening (NBS) to incorporate new methods for aggregating clinical and genomic data so that newer disease conditions can be added to the Recommended Uniform Screening Panel (RUSP) (20); 5) incentivizing compliance through financial and regulatory benefits that allow using real-world evidence (RWE) or pragmatic studies for RD clinical research; and 6) incentivizing research that includes MUPs by offering tax incentives to institutions that focus on recruiting these participants.

#### Pillar 3: Enforcement and governmental oversight

The US federal, state, and local government executive branches play a key role in enforcing and implementing DEIA mandates defined in relevant legislation, irrespective of the funding sources of RD research. We recommend that these governmental agencies 1) implement the mandated SOGI and SF data elements across clinical trial registries, patient registries, and any research programs managed by those agencies; 2) require demographics, including SOGI and SF results in research reporting stages; 3) establish a task force to oversee the implementation of complete, clear, accurate, and standardized collection and reporting of demographics and SF data to enhance interoperability, efficient decision-making, and to improve health equity for all; 4) support workforce development by encouraging existing federally funded programs to prioritize training and development opportunities in healthcare for those from MUPs backgrounds; 5) such as the FDA can consider updating the DAP 2024 guidance to include SOGI and SF as measures of diversity; and 6) such as the NIH and FDA could create guidelines for reporting demographics and SF data in publications.

#### Pillar 4: Implementation of standardized data collection, reporting, and publication

We recommend that stakeholders involved in conducting or reporting clinical trials 1) update or develop standard operating procedures for the complete, clear, accurate, and standardized collecting and reporting of demographics, including SOGI and SF by collaborating with international consortia such as the Clinical Data Interchange Standards Consortium (CDISC) and the International Council for Harmonisation (ICH) of Technical Requirements for Pharmaceuticals for Human Use; 2) such as publishers, journals, and editors should require authors to report demographics, including SOGI and SF data in published works or provide justifications if specific data were not reported; and 3) provide training, resources, and support for researchers to implement these reporting practices.

#### Pillar 5: Technological enablement

Inclusion and accessibility remain key challenges for RD patients. Technological advances in digital health, remote patient monitoring, virtual trials, and decentralized trials are transforming RD research and trials. We recommend 1) implementing decentralized and hybrid trial designs that leverage both randomized controlled trials and pragmatic, real-world data (RWD) that integrate remote monitoring and tele-visits where in-person visits are not justifiable; 2) using wearable technology to facilitate automated and continuous data collection from participants to minimize burden, especially for women, children, and older adults; 3) using artificial intelligence/ machine learning and natural language processing approaches to identify potential trial participants by mining RWD, such as EHRs, insurance claims, prescription drugs data, and registries, thereby minimizing bias in participant selection; 4) providing access to stable internet or digital devices for participants lacking these resources; and 5) implementing multichannel communication (audio, video, text, and multimedia) to accommodate those with special needs.

#### Pillar 6: Global epidemiology-based research

Historically, 90% of the clinical trials are conducted in 5% of the countries (Global North), excluding most low-and-middle-income countries. Each RD is a global public health issue with varying prevalence in different geographies, races, genetic haplogroups, and ethnicities. We recommend 1) supporting the understanding of epidemiological data such as prevalence and incidence of RDs in different geographic locations; 2) requiring trial designs that reflect the diversity of the populations affected by RDs globally; 3) expanding FDA’s involvement in global clinical trial networks to enable US sponsors access to the relevant global, diverse populations; and 4) agencies such as the NIH and FDA should require sponsors and clinical researchers to include epidemiological data as part of patient recruitment plans.

## Discussion

The findings of our study underscore the urgent need to improve MUP representation in RD clinical research. Therefore, we have proposed three specific recommendations and a six-pillar actionable framework (Tables 2 and 3, Figure 1). They broadly align with those of NASEM, the MRCT Center’s Diversity Guidance, the DCP, and the ASH DEI Toolkit (7–10). The key areas of overlap are standardized data collection and reporting, patient advocacy and engagement, improving access through technology, and governmental oversight. Our recommendation of including SOGI and SF as measures of diversity is aligned with the 21^st^ Century Cures Act, and FDORA (14,16). Building on these, we also recommend collecting and reporting special status, such as veteran, migrant, caregiver, and disability status, as these can influence individuals’ socioeconomic status and ability to access clinical trials.

Our framework proposes policy legislation as an essential pillar to achieving equitable representation of MUPs in RD clinical research. Our analysis demonstrated a clear association between the demographic variables reported in RD clinical research and the existing policy requirements (Table 2). While both the DCP and MRCT’s guidance acknowledge existing regulatory guidelines concerning clinical trial diversity, our framework considers legislative changes crucial for standardized data collection and reporting changes and improving inclusion and access to clinical trials. The Orphan Drug Act of 1983, a patient-led legislative initiative that spurred a revolution in developing treatments for RDs, exemplifies the impact of policy legislation (21). Modernizing NBS and expediting the implementation of the ICD-11 are crucial to enable patient participation in RD clinical research. Currently, the RUSP covers 35 core and 26 secondary conditions, most of which are RDs (20). Modernizing NBS can help reduce the time needed to diagnose RDs. The US healthcare system is yet to implement the ICD-11 codes, which covers about 5,500 unique RDs and synonyms (19). The ICD-10 system has specific codes for only 500 RDs. Adopting the ICD-11 can aid in data-driven monitoring of prevalence and incidences in MUP for at least half the known RDs. The global epidemiology pillar recognizes that each RD is a global public health challenge and acknowledges the geographic dispersion of affected individuals. US legislators have already recognized the need to modernize clinical trials through decentralized trials and are augmenting clinical trials with RWE (22,23). Given the challenges of RDs, our framework acknowledges the need to balance the urgency of making RD treatments available with the imperative of ensuring that research is diverse and inclusive. Hybrid trials and RWE can alleviate these difficulties. Therefore, we propose advocating for regulatory relaxation to allow the use of real-world evidence in regulatory decision-making.

Additionally, we want to note that a strong synergy between PAGs and legislators is required to bring sustainable changes to clinical research legislation. About 30% of the RD patient advocacy groups advocate for research funding, resources, and policies (24). PAGs and multi-stakeholder clinical research communities should educate lawmakers about the need for representative clinical research, including its economic benefits.

Several existing RD research, advocacy, and data-sharing initiatives (25–30) overlap with key components of our framework (Additional File 2: Table S1), highlighting its practical application. CDISC data standards for clinical trials have been successfully applied or recommended across regulatory agencies worldwide. Although the framework is aimed at RD clinical research, most of the recommendations can also be applied to other areas of clinical research where the underrepresentation of MUPs is prevalent (31–34).

While our framework and recommendations broadly align with existing frameworks and initiatives, there could be potential implementation barriers. Participants may have privacy and confidentiality concerns about collecting and reporting SOGI and intersectional identity data. Data security and confidentiality were reported as the main concerns about sharing health information in the database in the Rare-X DEI scoping review (35). These concerns should be addressed at the outset of participant engagement through linguistically diverse and culturally sensitive informed consent forms and materials that provide adequate information about the importance of these data and measures taken to safeguard them.

Although DEIA metrics can be standardized for the US, harmonized global minimum metrics are essential given the dispersed nature of people affected by RDs. For instance, while race and ethnicity are standard variables in clinical research in the US, countries like France and Germany do not collect ethnicity data (36). Therefore, these metrics must be adaptable to regional and sociocultural differences. Greater collaboration is required between international organizations like the CDISC, ICH, and regional regulatory bodies to align core DEIA metrics with add-on adaptable metrics for flexibility.

Technological advancements in streamlining clinical research with AI agents such as Jeeva Clinical Trials, to simplify, standardize, and automate the human workflows (37), can help reduce trial accessibility barriers for all stakeholders. This is especially true given the cost and time required to find patients meeting inclusion/exclusion criteria, especially for ultra-rare conditions. However, sponsors and trial sites should address the varying levels of comfort with technology among participants, the issue of 21% of the US population lacking home broadband connections (38), and issues of data and algorithmic biases in AI/ML (39,40) to further minimize the digital divide in clinical research participation.

Representative RD research is critical for scientific validity, especially given its low prevalence. Beyond the ethical and moral responsibility of making clinical research representative, stakeholders have a substantial economic incentive to embrace DEIA initiatives. The economic burden of RDs in the US alone is $997 billion (41). A recent NASEM report using the Future Elderly Model simulation projected potential gains of $33 trillion when racial and ethnic disparities were eliminated in chronic diseases through increased life-expectancy, disability-free years, and workforce participation (42). The study highlights the consequences of removing disparities for MUPs, suggesting that similar economic and public health benefits can be achieved if RD trials are made equitable and inclusive.

Our evaluation of policies, laws, and guidelines governing clinical research and suggested enhancements aim for holistic changes to the clinical research landscape. To illustrate, revising the Declaration of Helsinki (43) to ensure that informed consent forms are accessible to people from diverse backgrounds and needs, including various linguistic backgrounds and individuals with disabilities, will significantly advance clinical research to be truly inclusive and diverse.

Embedding DEIA principles in all documents governing clinical research can change the fabric of clinical research, bringing about a paradigm shift in clinical trial conduct and leading to equitable health outcomes for all.

### Limitations

Although our study could draw key inferences, it has certain limitations. First, the use of arbitrary cut-offs (low, moderate, high) for evaluating reporting statistics may limit the representation analysis. These cut-offs were based on the ideal of 100% reporting of variables to reveal broad trends in MUP representation. Second, considering participant distribution alone can limit our understanding of inclusion. Other factors such as eligibility criteria, accessibility barriers, and disease prevalence should also be considered for the complete picture. Third, since all RD trials were analyzed, comparing participant distribution with the US census was a practical choice to assess equity. Because RDs may disproportionately affect different populations, prevalence should be used as a benchmark for each RD. Fourth, our analysis was not stratified by the age-based prevalence of RDs, therefore insights regarding equity and inclusion can be distorted. Fifth, accessibility was inferred and could not be measured based on the reporting or distribution statistics, thus limiting the ability to quantify access challenges for MUPs with RD. Lastly, evaluating genetic ancestry as a diversity variable fell outside the scope of this work; however, it is worth noting that 80% of RDs are genetic in origin.

## Conclusions

This study is the first to evaluate the DEIA landscape in RD clinical research and policy and propose a framework to improve the participation of MUPs. Each pillar of our framework strengthens the foundation of RD clinical research through patient advocacy, policies, guidelines, implementation, enablement, and global impact, improving the representation of MUPs. We call upon patient advocacy groups, lawmakers, federal agencies, regulatory agencies, trial sites, trial coordinators, sponsors, industry, and researchers to adopt this framework for enhancing MUP participation in RD clinical research. Now is the time to take action for diverse, inclusive, equitable, accessible, and globally impactful RD research.

## Data Availability

The data used in this manuscript are derived from our companion manuscript, Manjunatha et al., Current State and Demographic Trends of Medically Underserved Populations in Rare Disease Research in the United States. Additional data may be available upon reasonable request.

## Abbreviations

MUPs: Medically underserved populations
DEIA: Diversity, Equity, Inclusion, Accessibility
RD: Rare disease
SF: Socioeconomic factors
US: United States
LGBTQ+: lesbian, gay, bisexual, transgender, queer or questioning
ASH: American Society of Hematology
DCP: Diversity Convergence Project
MRCT: Multi-Regional Clinical Trials
NASEM: National Academy of Science, Engineering, and Medicine
EHR: electronic health records
DAP: Diversity Action Plan
RD: Rare Disease
FDA: Food and Drug Administration
NIH: National Institute of Health
FDORA: Food and Drug Omnibus Reform Act
SOGI: Sexual Orientation and Gender Identity
PAG: Patient advocacy group
ICD: International Classification of Diseases
NBS: Newborn Screening
RUSP: Recommended Uniform Screening
RWE: Real-world evidence
CDISC: Clinical Data Interchange Standards Consortium
ICH: International Council for Harmonisation
RWD: Real-world data
ML: Machine learning
DEI: Diversity, Equity, Inclusion

## Declarations

### Ethics approval and consent to participate

Not applicable

### Consent for publication

Not applicable

### Availability of data and material

The data used in this manuscript are derived from our companion manuscript, Manjunatha et al., Current State and Demographic Trends of Medically Underserved Populations in Rare Disease Research in the United States, submitted to *BMC Health Services Research* (Submission ID 86faf02b-24c8-40c7-bdb4-1d1e06ba2e58). Additional data may be available upon reasonable request.

### Author contributions

HKR, LM, and SMS contributed to the conceptualization of the study. LM and SMS acquired data, developed methods, conducted investigation, validated the data, drafted the manuscript, and prepared figures; LM and SMS analyzed the data. LM, SMS, NV, RVK, HKR, JNW, and LGB interpreted the data, reviewed it for important intellectual content, and revised and edited the manuscript. HKR, NV, and RVK obtained funding for the study. LM managed the project administration with JNW and LGB, providing administrative, technical, or material support. HKR supervised the study. All authors read and approved the manuscript.

## Acknowledgments

IndoUSrare acknowledges the RDDC for providing funding. The authors thank the RDDC Clinical Trials Working Group members for their invaluable contributions during manuscript development. We acknowledge Dr Dora Akinyi Mugambi, Ms Leanne Marie Woehlke, Ms Kathleen Machuzak, Ms Veronica Mullins, and Dr Paul I Howard for analysis, interpretation, and content review and Ms Jocelyn Cooper and Ms Lolita Smith-Moore for administrative support. We also acknowledge support from the Children’s National Hospital.

## Authors’ information

Indo US Organization for Rare Diseases, Herndon, VA 20171, USA

Lavanyaa Manjunatha, PhD; Saundarya MS, MSc; Deepika Dokuru, PhD; Nisha Venugopal, PhD; Reena V Kartha, PhD; Harsha K Rajasimha MS, PhD

Center for Orphan Drug Research, Department of Experimental and Clinical Pharmacology, University of Minnesota Twin Cities, Minneapolis, MN, USA

Reena V Kartha, PhD

Jeeva Clinical Trials Inc., Manassas, VA 20109, USA; School of Systems Biology, George Mason University, Fairfax, VA 22030, USA

Harsha K Rajasimha MS, PhD

Rare Disease Diversity Coalition, Black Women’s Health Imperative, Atlanta, GA 3013, USA Jenifer Ngo Waldrop, MS; Linda Goler Blount, MPH

## Additional information

**Additional File 1_Manjunatha et al_Framework Inclusive and Accessible_RD Clinical Research_20032025.xlsx**

Additional File 1: Table S1.

**Additional File 2_Manjunatha et al_Framework Inclusive and Accessible_RD Clinical Research_06052025.docx**

Additional File 2: Supplementary methods, Table S1: Existing programs or initiatives that overlap with our framework

