## Supplementary Methods and Table for "A Framework for Inclusive and Accessible Clinical Research in Rare Diseases"

### Representation gaps analysis

Data from the companion manuscript and supplementary tables (1) were extracted for the 13 variables. Ideally, the percentage of publications or trials reporting any demographic or socioeconomic factor (SF) variable should be 100%. In this analysis, categorization (high: >75%, moderate: 25–75%, low: <25%) of reporting statistics was arbitrary and was chosen to facilitate the interpretation of the data in the absence of standardized guidelines. The same categorization was applied to participant distribution statistics. However, the equity analysis was based on comparing with the US census. Reporting statistics were evaluated based on the percentage of publications or trials (high: >75%, moderate: 25–75%, low: <25%) reporting a specific variable and its subcategories. Representation was evaluated based on whether data were available for all subcategories of a variable and levels of reporting. The distribution statistics were evaluated based on the proportion of participants (high: >75%, moderate: 25–75%, low: <25%) in each subcategory of a variable. Finally, participant distributions were compared to the US census through representation ratios calculated by dividing the proportion of participants in the trial by the proportion in the US census (2). Representation ratios >1 indicated that a group was well represented, and ratios <1 indicated underrepresentation. We did not calculate representation ratios for the age variable as three of its six subcategories included participants of mixed age groups, therefore not allowing for a meaningful direct comparison with the US census.

### Policy analysis

Google searches were performed using the search terms “US policies for clinical research,” “Federal guidelines and policies for clinical research,” and “US legislation clinical trials.” On the Congress.gov website, the queries “diversity clinical trials” and “clinical trials” were searched after applying the filter Subject - Policy Area = Health. The FDA guidance documents list was downloaded from and filtered by guidance title and topic using the terms “diversity,” “race,” “ethnicity,” and “postmarketing surveillance.” All resulting searches were first screened by title and content. Only those documents directly addressing diversity in clinical trials, participant protections, informed consent, and demographic data submission were included.

### Table S1: Existing programs or initiatives that overlap with our framework

| **Program or initiative name** | **Description** | **Overlaps with  our framework** |
| --- | --- | --- |
| IAMRARE program (3) | A patient-registry platform run by the National Organization for Rare Diseases | Patient Advocacy |
| Rare Disease Cures Accelerator-Data Analytics Program (4) | An FDA-funded initiative to curate and standardize data from diverse sources to accelerate rare disease characterization and treatment development | Standardized data collection and sharing  Technological enablement |
| Rare Disease Legislative Advocacy Program (5) | Supports the legislative advocacy of all RD patients and organizations | Patient and Legislative advocacy |
| Rare Disease International (6) | A global alliance of patients with rare disease advocating for rare diseases as a global health priority | Global legislative or policy advocacy  Technological enablement |
| Rare-X (7) | A collaborative global data-sharing platform to accelerate rare disease treatments, focusing on DEI through patient-stewarded data collection and other initiatives like the DEI scoping review project (8) | Global-epidemiology  Standardized data sharing  General DEI focus |
| Rare Disease Diversity Coalition (9) | Dedicated to addressing the extraordinary challenges faced by historically underrepresented rare disease patients as encompassed by social determinants of health | Patient advocacy  Legislative advocacy/policy legislation |
| Indo US Organization for Rare Diseases (10) | Nonprofit organization promoting rare disease research, patient and policy advocacy by facilitating collaboration between stakeholders in India and the US. | Patient advocacy, Legislative advocacy/policy legislation  Global epidemiology |

Abbreviation: DEI, diversity, equity, inclusion; US, United States
